# An Aetiological and Observational Cohort study of Interstitial Lung Abnormalities (ILAs) Including Progressive Fibrosing Phenotype Interstitial Lung Diseases in a Large General Medical Check-up Population (Kumamoto ILA study in Japan) (KILA-J)

**DOI:** 10.1101/2023.01.04.23284128

**Authors:** Kazuya Ichikado, Hidenori Ichiyasu, Kazuhiro Iyonaga, Kodai Kawamura, Noritaka Higashi, Takeshi Johkoh, Kiminori Fujimoto, Jun Morinaga, Minoru Yoshida, Katsuhiko Mitsuzaki, Moritaka Suga, Naoya Tanabe, Tomohiro Handa, Toyohiro Hirai, Takuro Sakagami

## Abstract

**Introduction:** Interstitial lung abnormalities (ILAs) are subtle or mild parenchymal abnormalities observed in more than 5 % of the lungs on CT scans in patients in whom interstitial lung disease was not previously clinically suspected and is considered. ILA is considered to be partly undeveloped stages of idiopathic pulmonary fibrosis (IPF) or progressive pulmonary fibrosis (PPF). This study aims to clarify the frequency of subsequent IPF or PPF diagnosis, the natural course from the preclinical status of the diseases, and the course after commencing treatment.

**Methods and analysis:** This is an ongoing, prospective, multicentre observational cohort study of patients with ILA referred from general health screening facilities with more than 70000 annual attendances. Up to 500 participants will be enrolled annually over 3 years, with 5-year assessments every six months. Treatment intervention including anti-fibrotic agents will be introduced in disease progression cases. The primary outcome is the frequency of subsequent IPF or PPF diagnoses. Additionally, secondary and further endpoints are associated with the efficacy of early therapeutic interventions in cases involving disease progression.

**Ethics and dissemination:** This study protocol and informed consent documents have been approved by the Institutional Review Boards of Kumamoto University Hospital (approval number: 2368), Saiseikai Kumamoto Hospital (approval number: 809), and each participating institution. Additionally, written informed consent will be obtained from all participants. Patient recruitment commenced on 20 June 2022. The results will be disseminated through peer-reviewed publications and international conferences.

**Trial registration number:** UMIN000045149

**Strengths and limitations of this study:** This is the first prospective, multicentre, observational study to clarify the following points:

- the aetiological data of patients with ILA from a large general health check-up population
- the natural course of IPF or PPF from the asymptomatic stage
- the effects and outcomes of early therapeutic intervention including anti-fibrotic agents for progressive cases of ILA.

The limitations of the study:

- participants missing the regular 6-monthly visits after consenting to participate in the study because of their asymptomatic or the pandemic viral infection.

## INTRODUCTION

Interstitial lung abnormalities (ILAs) are defined as incidental computed tomography (CT) findings of non-dependent abnormalities affecting more than 5% of the lungs (1-3). The clinical importance of ILAs has been increasingly recognised as early and asymptomatic lesions in cases of progressive fibrosing interstitial lung diseases (ILDs) (4,5). Furthermore, the presence of ILAs is a prognostic factor in ILD, lung cancer, and chronic obstructive pulmonary disease (COPD). This is because ILAs are subcategorised as non-fibrotic and fibrotic, with fibrotic abnormalities thought to represent early or mild pulmonary fibrosis, resulting in a higher risk of complications in lung cancer or COPD (1-3,6). Although an asymptomatic period in which ILAs can be detected during the high-resolution CT (HRCT) examinations is likely to precede a pulmonary fibrosis diagnosis, the natural history of progression from ILA stage to overt disease remains unknown (4,5). This challenge has remained unresolved with progressive pulmonary fibrosis (PPF) as a newly defined pathological concept characterised by progressive fibrosis as well as IPF (5,7). Furthermore, the effect of early anti-fibrotic drug therapies for ILA on progressed disease stages is unknown. However, anti-fibrotic drugs have effectively prevented the decline of forced vital capacity (FVC) with a potential for improved IPF or PPF prognoses (5,7).

The frequency of ILA and predictors of ILA progression have been identified in large studies on COPD cases and populations at risk of cardiovascular diseases (3,6), however, there is limited data on ILA in many general check-up populations. Hence, this study aims to clarify the natural course from ILA or the asymptomatic stage in patients subsequently diagnosed with IPF or PPF and the course and outcome after the introduction of therapeutic interventions. Additionally, this study also aims to clarify the frequency and progression index of ILA by analysing ILA cases screened from general health screening facilities with the largest number of examinees.

## METHODS AND ANALYSIS

### Study design

This is a prospective, multicentre observational cohort study on ILAs diagnosed by respiratory specialists from general health screening facilities in Kumamoto Prefecture, Japan. Other details of the study’s design are as follows:

- Presence or absence of invasion: Mild invasion (no new needle insertion with plasma blood sampling at enrollment:)
- Presence or absence of intervention: No intervention
- Presence or absence of use of samples collected from the human body: Yes (plasma and serum are stored)
- Positioning of the study: observational study
- Randomization: None

### Participant selection and screening

#### Study population

Training sessions will be held for the physicians in the screening facilities participating in the study to confirm the study participants using any of the following three findings, (i) the presence of fine crackles on auscultation, (ii) opacification of the costophrenic angles on chest X-ray, or (iii) the presence of subpleural ground-glass or reticular shadows on CT findings. Cases with either of these findings in the screening facilities are suspected cases of ILA. The study will enroll approximately 500 patients from >70000 examinees in general health check-up facilities, annually screened by chest radiography, with fine crackles on auscultation and/or computed tomography findings. Participants will be enrolled over 3 years and followed up for 5 years. The primary outcome is the evaluation of the aetiological data of preclinical patients who are subsequently diagnosed with IPF or PPF. This study has been designed according to the Strengthening the Reporting of Observational Studies in Epidemiology Statement guidelines. Patients or the public were involved in the design, or conduct, or reporting, or dissemination plans of our research.

#### Inclusion Criteria

Patients with one or more of the following findings during check-ups will be suspected of having ILAs which is slightly modified in the Fleiscner Society criteia (1) : (i) fine reticular shadows at the costophrenic angles on chest radiography, (ii) fine crackles on auscultation, and (iii) lower predominant subpleural reticular shadows on CT findings. Patients suspected to have ILAs at a screening facility will be referred for further examination in any of the three teaching hospitals (Kumamoto University Hospital, Japanese Red Cross Kumamoto Hospital, and Saiseikai Kumamoto Hospital) where pulmonologists work. Patients diagnosed with ILAs confirmed using HRCT who provided informed consent will be enrolled into the study’s database.

This study defines ILAs as subtle or mild parenchymal abnormalities observed in >5% of the lungs during CT examination of patients in whom ILD has not been previously clinically suspected.

#### Exclusion Criteria

The following are exclusion criteria: (i) patients who refuse to participate in the study after reading the explanation, (ii) patients with acute or subacute progressive interstitial pneumonia, (iii) patients who have difficulties undergoing the periodic follow-up due to their distant places of residence, (iv) cases that are assessed as inappropriate for the study by medical attendants, (v) cases incorporated into intervention studies, such as clinical trials that are not allowed to participate in observational studies.

#### Data collection and management

##### Primary endpoint

The primary endpoint is the frequency of subsequent IPF or PPF diagnoses.

##### Secondary endpoints

The secondary endpoints are (i) frequency of ILA cases among all general health examiners and (ii) frequency of fibrotic hypersensitivity pneumonitis and collagen vascular diseases related to interstitial pneumonia in patients diagnosed with PPF.

##### Further endpoints

The progression rate of ILA cases will be examined using HRCT. In addition, IPF cases involving patients with ILA will be evaluated for: (i) the rates of decline in FVC, percentage diffusing capacity for carbon monoxide (%DLco), and 6-minute walk distance; and (ii) the progression rate of the ILD-Gender–Age–Physiology (ILD-GAP) GAP Index. Rate of decline in 6-minute walk distance in cases of IPF diagnosed from ILA patients The progression rate of HRCT findings in IPF cases involving patients with ILA will be assessed by comparing the visual and quantitative assessments using artificial intelligence-based quantitative CT (AIQCT) (12). In addition, the progression rate of HRCT findings in PPF cases involving patients with ILA will be assessed by comparing the visual and quantitative assessments using AIQCT. Furthermore, the proportion of HRCT findings showing progression in IPF or PPF cases before and after therapeutic interventions will be assessed by comparing the visual and quantitative assessments using AIQCT.

Other endpoints to be assessed include: (i) the association between HRCT progression indices and functional indices or clinical outcomes in patients with IPF or PPF, (ii) the rate of FVC decline before and after anti-fibrotic drug therapy in IPF and PPF cases involving patients with ILA, (iii) mortality rates of IPF from the asymptomatic stage, (iv) incidence of acute exacerbation in IPF and PPF cases from the asymptomatic stage, (v) the frequency of respiratory complications (pneumonia, lung cancer, and others) requiring inpatient treatment of patients with IPF or PPF from the asymptomatic stage, (vi) rate of decline in %DLco before and after therapeutic intervention in PPF cases involving patients with ILA, and (vii) the output score of image AI to assess suspicions of interstitial pneumonia from chest X-rays (13).

#### Data Collection

There is insufficient evidence to support the cost-benefit ratio of screening asymptomatic individuals. Nonetheless, this study’s testing plan follows the recommendations of the Fleischner Society Position Paper 2020 (1). Details of data that will be collected during the scheme are described in the Online Supplementary Materials (Supplementary Table 1). Patients will be enrolled after confirming that they fulfill all inclusion criteria without meeting any exclusion criteria and they provide informed signed consent. In addition, the following data will be collected and registered at enrolment: (i) sex, date of birth, height, weight, and body mass index; (ii) reason for a referral from screening facilities; (iii) smoking history, environmental history (presence or absence of dust exposure, contact with birds, and occupational history), and family history of interstitial pneumonia; (iv) comorbidities, including diabetes mellitus, coronary artery disease, gastroesophageal reflux, cardiovascular diseases other than coronary artery disease, and lung cancer; and (v) presence of fine crackles on auscultation, chest radiography, and high-resolution CT findings and patterns based on international guidelines (7).

The following physical examination results will be collected every six months: oxygen saturation (SpO_2_) levels and dyspnoea scale score (modified Medical Research Council), pulmonary function tests, blood gas analysis, and a 6-minute walk test. Other tests with results include biological assessments (including blood count and renal and hepatic functions), autoantibody screening blood sampling, serum interstitial pneumonia markers (Krebs von den Lungen-6 (KL-6), surfactant protein-D (SP-D)), ILD-GAP index, and echocardiography results. Suspected diagnosis and diagnostic accuracy of IPF based on ontology will be recorded after discussing the multidisciplinary diagnosis (MDD) via a web conference involving the three institutions (MDD diagnosis) and each institution’s diagnosis.

Patients that show any signs of disease progression, including the appearance of clinical symptoms, decline in lung function, or exacerbation of HRCT findings evaluated by visual assessment and AIQCT (12), will be considered for a full work-up and the introduction of drug therapy. The detailed examination will include bronchoalveolar lavage (BAL), transbronchial lung cryobiopsy, or surgical lung biopsy. After registration, subsequent pulmonary complications requiring hospitalisation, such as acute exacerbation, pneumonia, pneumothorax, or lung cancer, will be recorded.

Furthermore, it will be recorded if long-term oxygen therapy is needed in cases of progression despite appropriate medication during the follow-up years. When therapeutic intervention is introduced, the assessment plan is reset, and regular assessments are carried out during the period from the introduction of treatment (Supplementary Table 2).

### Management of enrolled participants

Data and images obtained during routine observation will be shared with the three medical centres via electronic data capture (EDC) and the imaging cloud. Based on the data, each of the three medical centres performs a case-specific institutional multidisciplinary discussion diagnosis (MDD) and registers it in the EDC. HRCT images will also be evaluated independently by two experienced chest radiologists (T. J. and K. F.), and the results will be entered into the EDC. Cases in which cryobiopsy or surgical lung biopsy is performed are referred to a pulmonary pathologist for pathological diagnosis, and the results are recorded on the EDC. In addition, cases registered at the three scrutinising medical centres are regularly web-case-conferenced between facilities. Furthermore, the facility MDD diagnosis and the web-conference MDD diagnosis will be recorded on the EDC.

### Statistical analysis

#### Sample size calculation

There are ≥70000 examinees yearly for two medical check-ups at health care centres (the Japanese Red Cross Kumamoto Hospital Health Care Center and the Center for Preventive Medicine of Saiseikai Kumamoto Hospital). The number of participants suspected of having ILAs was approximately 300 at each centre in 2018. Therefore, the total number of participants with ILAs is assumed to be approximately 600 per year.

According to the previous data, the number of cases will be underestimated by 10% because approximately 10% of the medical check-ups that require detailed tests for ILAs did not undergo the subsequent detailed examinations. Therefore, the maximum number of suspected ILAs is expected to be 500 cases annually.

Continuous variables will be expressed as medians and interquartile ranges (IQRs), and categorical variables for each group will be expressed as the number of subjects and percentages. The inter-observer variation in the presence or absence of HRCT findings will be analysed using the weighted kappa statistic. Inter-observer variation regarding the extent of the HRCT findings was assessed using Spearman’s rank correlation coefficient. The HRCT scores assigned by the two independent observers are compared using the Bland–Altman method. If the HRCT patterns or scores do not agree between the two radiologists; one of the patterns or scores is adopted by consensus.

All analyses with ILA progression as the outcome variable are performed using logistic regression, where ILA progression was dichotomised with progression defined as probable or definite progression, and regression defined as no change, probable regression, and definite regression using subjective visual estimation as well as an objective quantitative AI assessment (AIQCT). Multivariable analyses will be adjusted for variables such as age, sex, and smoking status. In addition, a Cox proportional-hazards model has been created to assess treatment efficacy, considering the immortal period as a time-dependent covariate. Furthermore, morbidity and mortality related to respiratory disease will be compared between groups using Chi-squared or Fisher’s exact tests. Finally, variation in the global extent of ILA on HRCT examination between each evaluation time point will be compared between groups using the Mann-Whitney U or Wilcoxon tests.

Clinical laboratory evaluations will be expressed as mean, median, and interquartile range for each evaluation time point, and boxplots will be created for graphical representation. In addition, percentages of abnormal clinical laboratory results will be compared between the groups using Chi-squared or Fisher’s exact tests. No interim data analysis has been planned.

## DISCUSSION

This is the first prospective, multicentre, observational study to clarify (i) the aetiological data of patients with ILA from a large general health check-up population, (ii) the natural course of IPF or PPF from the asymptomatic stage, and (iii) the effects and outcomes of early therapeutic intervention including anti-fibrotic agents for progressive cases of ILA.

### Aetiological data of patients with ILA from the general health check-up population

The AGES-Reykavik study, an epidemiological study including patients aged ≥40 years at risk of cardiovascular lesions, reported that 378 (7%) patients with ILAs on initial HRCT were observed from 5320 participants. Furthermore, 327 of 378 patients who underwent subsequent CT scans were evaluated for ILA progression (6). The AGES-Reykavik study reported that 238 (73%) of 327 participants eventually showed ILA progression over 5 years (median: 5.1 years). ILA progression was also assessed in 1867 participants in the Framingham Heart Study (6) using serial CT scans. The study reported that 155 (8%) of 1867 participants showed ILAs on initial CT scans. Furthermore, 119 (76%) of the 155 participants showed ILA progression on serial CT scans during the follow-up period (6 years).

Although both studies did not describe the specific diagnoses in cases that showed ILA progression, the mortality rate from pulmonary fibrosis in the group with ILA was reported to be as high as 47% in the AGES-Reykjavik study.

The present clinical study is supposed to include a large number of participants with ILAs observed on HRCT, especially in the largest population (>210000 examinees) of general health check-ups. Hence, the study’s primary and further endpoints will be achieved.

### Specific MDD diagnosis for cases progressing from the ILA stage and response to early therapeutic intervention

It has been reported that 20% of ILA cases progress within 2 years, and >40% progress within 5 years (1,2). However, there has been little data about how many patients are subsequently diagnosed with IPF or PPF. Furthermore, the natural history of the asymptomatic phases corresponding to the ILA stages in these progressive fibrosing phenotypes has not been elucidated. A recent meta-analysis showed that anti-fibrotic treatment reduces the risk of all-cause mortality in progressive fibrosing phenotypes, including IPF (8,9). However, it is critical to determine whether the early identification and diagnosis of these fibrotic ILDs could lead to early treatment, resulting in a better prognosis (5). The present study’s flow is to assess the ILA stage every 6 months. Once clinical progression is confirmed, a full examination will be considered, including BAL and biopsy, according to the 2022 International Guidelines, and the degree of confidence in MDD diagnosis. Clarification on the progression from ILA to an advanced disease stage and treatment responsiveness in the respective pathologies of IPF and PPF is expected.

### Indicators of functional impairment during the ILA phase

Functionally, reductions in total lung capacity and reduced exercise capacities, such as a greater reduction in SpO_2_ values during the 6-minute walk test or 6-minute walk distance, have been reported in the ILA group compared with those in the non-ILA group (10). However, there are no data on how often functional impairments should be assessed during the ILA phase. Although differences in ILA prognosis have been reported according to different initial HRCT patterns (11), differences in the progression of functional disabilities are also expected. Indicators of functional impairment during the ILA phase have not been fully investigated according to the initial HRCT patterns of the ILA. The present study hopes to clarify whether these functional test abnormalities in the ILA phase are observed at 6-month test intervals and whether there are differences in HRCT imaging patterns.

### Relationship between artificial intelligence analysis for quantification of ILA on HRCT scans and clinical outcomes

Artificial intelligence and machine learning approaches have become increasingly important for quantifying and classifying ILA and fibrotic ILDs (12). They will also help address the increasing demand for identifying the progression of ILA. In this study, quantitative assessments of ILA using AI will be combined with visual assessments that semi-quantify the visual extent of ILA in 5% increments, as well as the pattern of HRCT findings in line with the 2022 IPF international guidelines. The presence or absence of progression will be assessed quantitatively using AI (AIQCT) and semi-quantitatively by visual assessment. The relationship between the degree of progression and HRCT pattern at enrolment and changes in lung function indices and serum markers will also be investigated.

### Study limitations

First, since almost all patients suspected to have ILAs are asymptomatic, they may not necessarily visit the teaching hospitals after referrals. For example, previous data from one of the check-up facilities (The Center for Preventive Medicine of Saiseikai Kumamoto Hospital) shows that the rate of missing re-evaluation was approximately 10%. Second, patients may fail to attend the regular 6-monthly visits after consenting to participate in the study because they are asymptomatic. Finally, a viral infection pandemic, such as COVID-19 or flu, might also affect regular visits every 6 months. For example, during the COVID-19 pandemic, patients feared the infection and tended to withhold regular hospital visits.

## Data Availability

Although this study has been ongoing now, all data produced in the study will be available upon reasonable request to the authors.

## Authors’ contributions

KIchi, HI, KIyo, KK, NH, TJ, KF, JM, MY, KM, NT, TH, MS, TH, and TS were all involved in the study conception and conducted, data analysis, and drafting or revision of the manuscript. Each author is accountable for the accuracy and integrity of the research output.

JM is a biostatistician and is especially in charge of statistical analysis.

## Patient and public involvement

Patients and/or the public were not involved in the design, conduct, reporting, or dissemination plans of this research.

## Patient consent for publication

Obtained.

## Provenance and peer review

Not commissioned; externally peer-reviewed.

**Supplementary Table 1.** Schedule of Activities

| Time<br>Activities | Entry<br>Period of time allowed | 6 Mo. | 12 Mo. | 18 Mo. | 24 Mo. | 30 Mo. | 36 Mo. | 42 Mo. | 48 Mo. | 54 Mo. | 60 Mo. | At the time of Discontinuation |
| --- | --- | --- | --- | --- | --- | --- | --- | --- | --- | --- | --- | --- |
|  |  | 3- 9 Mo. | 9- 15 Mo. | 15- 21 Mo. | 21- 27 Mo. | 27- 33 Mo. | 33- 39 Mo. | 39- 45 Mo. | 45- 51 Mo. | 51- 57 Mo. | 57- 63 Mo. |  |
| Consent | ○ | — | — | — | — | — | — | — | — | — | — | — |
| Patient background (age, date of birth, gender, height, weight) | ○ | — | — | — | — | — | — | — | — | — | — | — |
| Reasons for referral from screening facilities | ○ | — | — | — | — | — | — | — | — | — | — | — |
| Family history of interstitial pneumonia | ○ | — | — | — | — | — | — | — | — | — | — | — |
| Smoking History (Brinkman Index) | ○ | — | — | — | — | — | — | — | — | — | — | — |
| Dust Exposure History | ○ | — | — | — | — | — | — | — | — | — | — | — |
| Occupational Art (organic solvents, organic dusts, inorganic dusts) | ○ | — | — | — | — | — | — | — | — | — | — | — |
| Comorbidities/pre-existing medical conditions (diabetes, coronary artery disease, cardiovascular diseases other than coronary artery disease, gastro-esophageal reflux) | ○ | — | — | — | — | — | — | — | — | — | — | — |
| Environmental history of bird-related exposure (e.g. history of duvet use, poultry manure, stuffed birds, aviaries) | ○ | — | — | — | — | — | — | — | — | — | — | — |
| Housing history (e.g. age, sun exposure, mould) | ○ | — | — | — | — | — | — | — | — | — | — | — |
| Chest X-P | ○ | ○ | ○ | ○ | ○ | ○ | ○ | ○ | ○ | ○ | ○ | △ |
| High-resolution CT | ○ | ○ | ○ | ○ | ○ | ○ | ○ | ○ | ○ | ○ | ○ | △ |
| HRCT pattern<br>UIP<br>Probable UIP<br>Indeterminate for UIP<br>Alternative diagnosis<br>ILA pattern | ○ | ○ | ○ | ○ | ○ | ○ | ○ | ○ | ○ | ○ | ○ | △ |
| HP guideline<br>HRCT pattern<br>Non-fibrotic | ○ | ○ | ○ | ○ | ○ | ○ | ○ | ○ | ○ | ○ | ○ | △ |
| Fibrotic<br>Typical<br>Compatible with HP<br>Indeterminate for HP |  |  |  |  |  |  |  |  |  |  |  |  |
| Extent of HRCT findings (visual<br>assessment: 5% increments ) | ○ | ○ | ○ | ○ | ○ | ○ | ○ | ○ | ○ | ○ | ○ | △ |
| Extent of HRCT findings<br>(assessed by AIQCT) | ○ | ○ | ○ | ○ | ○ | ○ | ○ | ○ | ○ | ○ | ○ | △ |
| Blood gas analysis | ○ | — | ○ | — | ○ | — | ○ | — | ○ | — | ○ | △ |
| 6MWT | ○ | △ | ○ | △ | ○ | △ | ○ | △ | ○ | △ | ○ | △ |
| PFT FVC | ○ | ○ | ○ | ○ | ○ | ○ | ○ | ○ | ○ | ○ | ○ | △ |
| %FVC | ○ | ○ | ○ | ○ | ○ | ○ | ○ | ○ | ○ | ○ | ○ | △ |
| VC | ○ | ○ | ○ | ○ | ○ | ○ | ○ | ○ | ○ | ○ | ○ | △ |
| %VC | ○ | ○ | ○ | ○ | ○ | ○ | ○ | ○ | ○ | ○ | ○ | △ |
| %DLco | ○ | ○ | ○ | ○ | ○ | ○ | ○ | ○ | ○ | ○ | ○ | △ |
| %DLco/VA | ○ | ○ | ○ | ○ | ○ | ○ | ○ | ○ | ○ | ○ | ○ | △ |
| FEV1.0 | ○ | ○ | ○ | ○ | ○ | ○ | ○ | ○ | ○ | ○ | ○ | △ |
| FEV1.0% | ○ | ○ | ○ | ○ | ○ | ○ | ○ | ○ | ○ | ○ | ○ | △ |
| Modified MRC | ○ | ○ | ○ | ○ | ○ | ○ | ○ | ○ | ○ | ○ | ○ | △ |
| Cough, presence of fine<br>crackles on auscultation | ○ | ○ | ○ | ○ | ○ | ○ | ○ | ○ | ○ | ○ | ○ | △ |
| Autoantibody screening blood<br>sampling | ○ | — | ○ | — | ○ | — | ○ | — | ○ | — | ○ | △ |
| Specific autoantibody blood<br>sampling | △ | — | △ | — | △ | — | △ | — | △ | — | △ | △ |
| Presence of symptoms of<br>connective tissue diseases | ○ | ○ | ○ | ○ | ○ | ○ | ○ | ○ | ○ | ○ | ○ | △ |
| Diagnosis of interstitial lung<br>disease | ○ | ○ | ○ | ○ | ○ | ○ | ○ | ○ | ○ | ○ | ○ | △ |
| ILD-GAP score · stage | ○ | ○ | ○ | ○ | ○ | ○ | ○ | ○ | ○ | ○ | ○ | △ |
| Diagnostic accuracy of IPF<br>(≤90%Confident, 89-70%<br>Provisional high confident, 69-50% Provisional low confident,<br>50%≥ Unclassifiable) | ○ | ○ | ○ | ○ | ○ | ○ | ○ | ○ | ○ | ○ | ○ | △ |
| ECG | ○ | — | — | — | — | — | — | — | — | — | ○ | △ |
| # blood biochemistry test | ○ | ○ | ○ | ○ | ○ | ○ | ○ | ○ | ○ | ○ | ○ | △ |
| Serum marker(KL-6, SP-D, ACE<br>(optional) , sIL-2R (optional) | ○ | ○ | ○ | ○ | ○ | ○ | ○ | ○ | ○ | ○ | ○ | △ |
| Stored plasma and serum<br>collection | ○ | — | — | — | — | — | — | — | — | — | ○ | △ |
| Bronchoalveolar lavage(BAL)<br>(optional) * | △ | △ | △ | △ | △ | △ | △ | △ | △ | △ | △ | △ |
| Transbronchial cryobiopsy<br>(optional) * | △ | △ | △ | △ | △ | △ | △ | △ | △ | △ | △ | △ |
| Surgical lung biopsy (optional) * | △ | △ | △ | △ | △ | △ | △ | △ | △ | △ | △ | △ |
| Presence or absence of acute exacerbations | — | ○ | ○ | ○ | ○ | ○ | ○ | ○ | ○ | ○ | ○ | ○ |
| New onset of pneumonia (infection) | — | ○ | ○ | ○ | ○ | ○ | ○ | ○ | ○ | ○ | ○ | ○ |
| Presence or absence of lung cancer complications | ○ | ○ | ○ | ○ | ○ | ○ | ○ | ○ | ○ | ○ | ○ | ○ |
| Presence of complications requiring inpatient treatment | — | ○ | ○ | ○ | ○ | ○ | ○ | ○ | ○ | ○ | ○ | ○ |
| Availability of home oxygen therapy | — | ○ | ○ | ○ | ○ | ○ | ○ | ○ | ○ | ○ | ○ | ○ |
| Reasons for discontinuation of the study/outcome | — | — | — | — | — | — | — | — | — | — | — | ○ |

**Supplementary Table 2.**
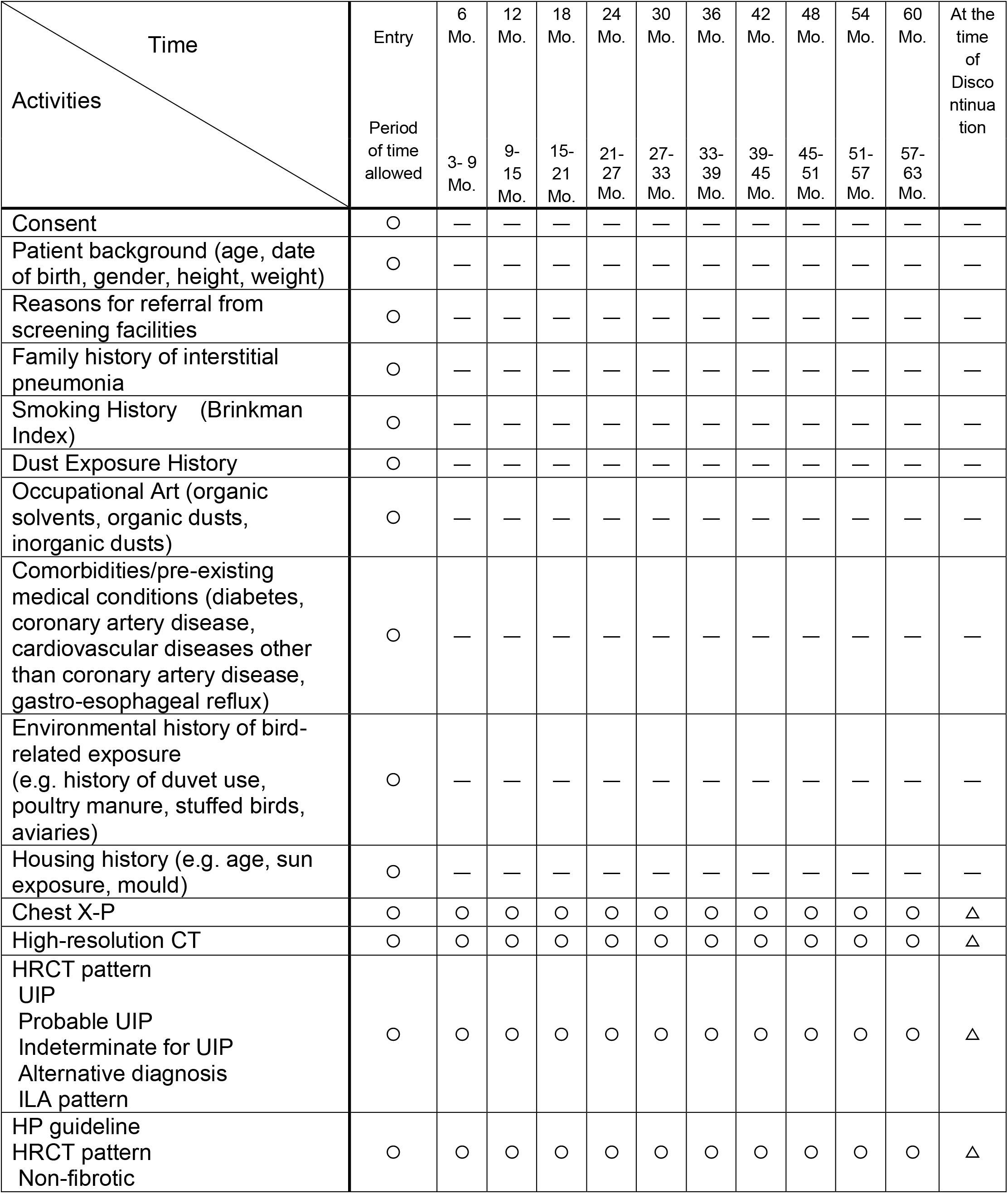
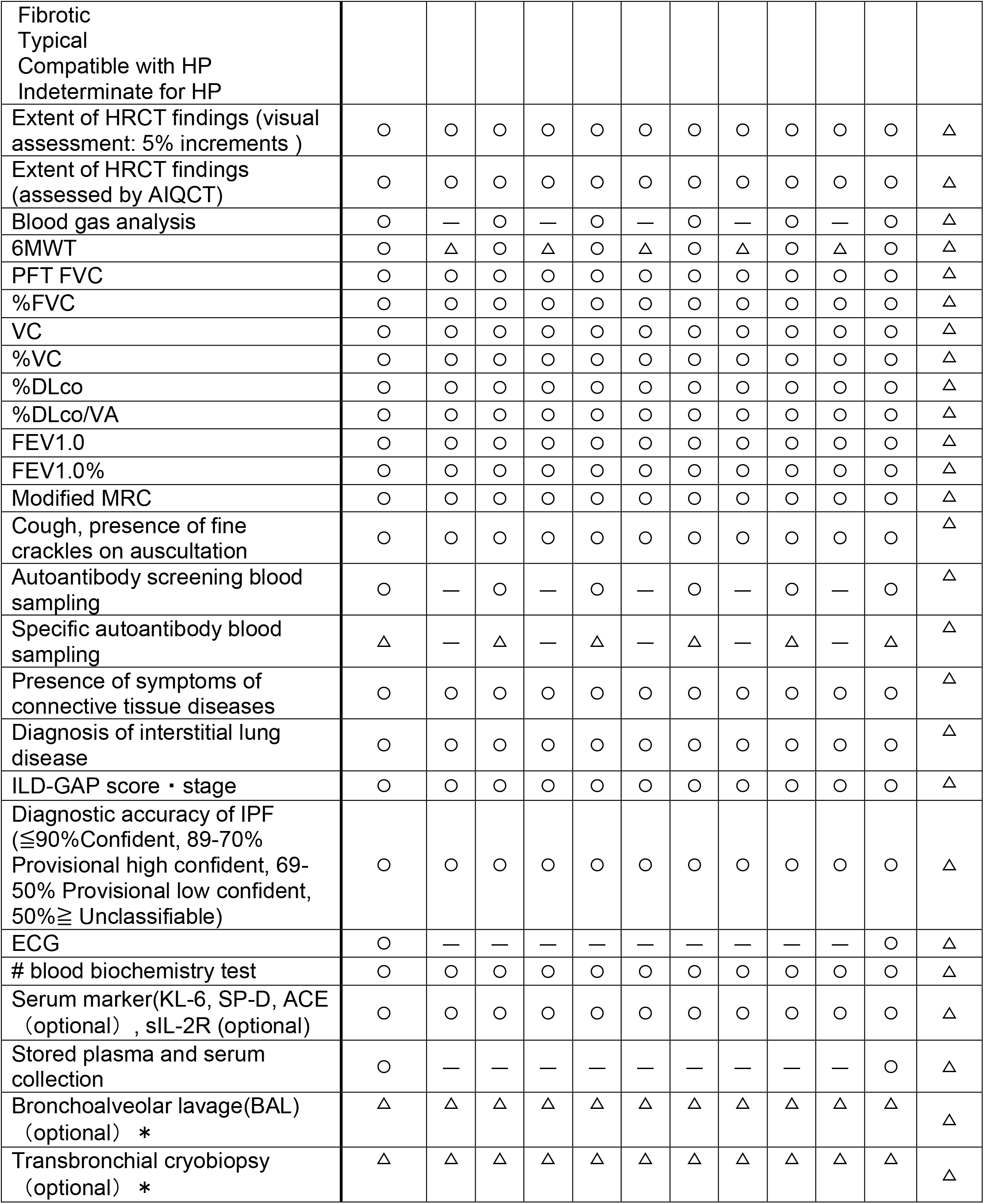
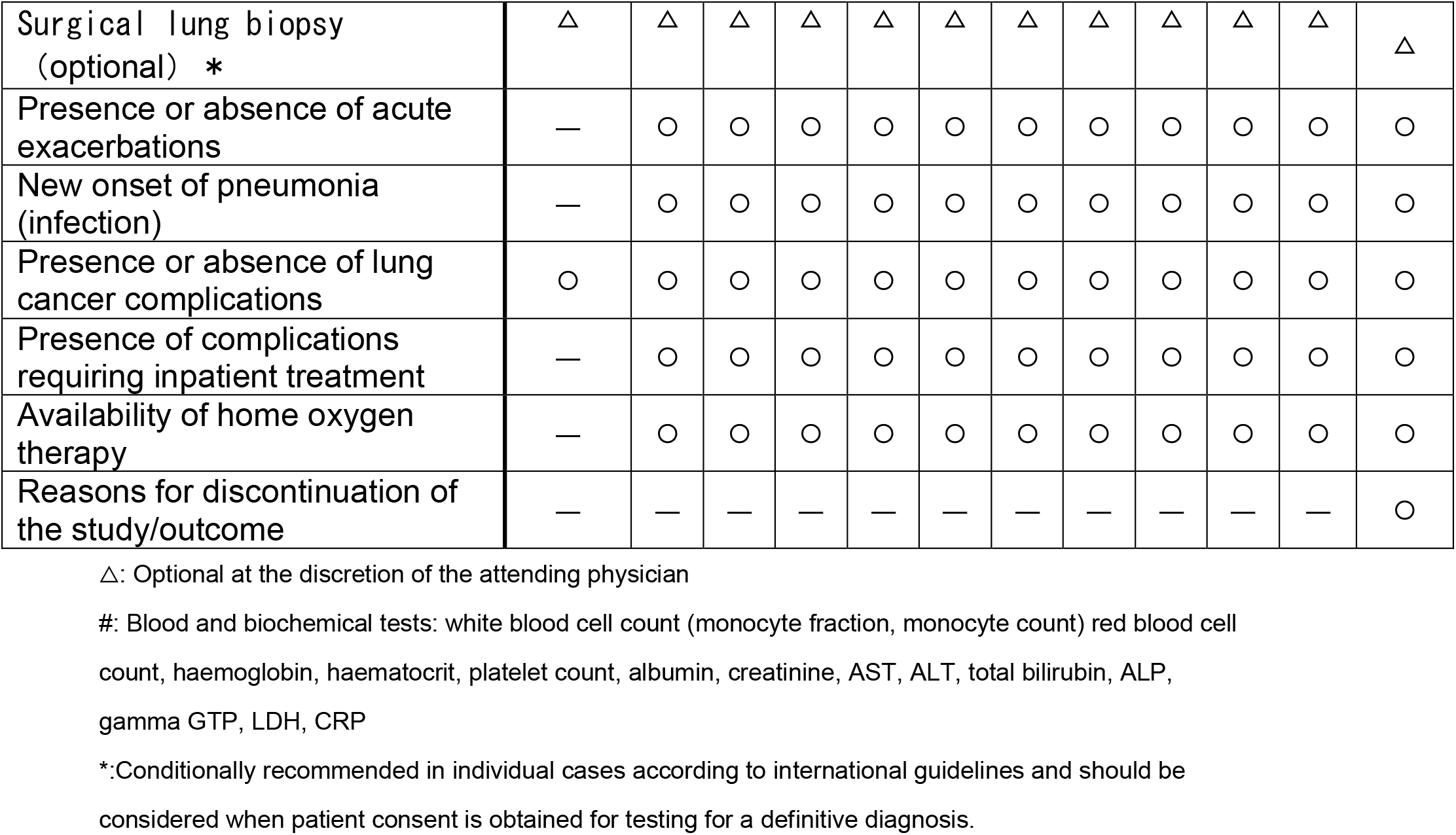
Schedule of Activities after introduction of treatment

